# A Novel Host Stool mRNA Extraction Method Improves Sensitivity of Detection for Colorectal Cancer and Precancerous Lesions

**DOI:** 10.64898/2026.01.20.25342587

**Authors:** Jakob Kirchner, Haijiu Lin, Loren Hansen, Janet Cheng, Changpu Song, Kristin Ramos, Dan Chen, Yanyang Liang, Wenying Pan

**Author notes:** Corresponding author Wenying Pan.

## Abstract

**BACKGROUND:** Early detection of colorectal cancer (CRC) is essential for reducing disease-related mortality; however, the invasiveness and resource intensity of colonoscopy limit its widespread use in population screening. Although several non-invasive screening modalities are available, their sensitivity remains suboptimal, particularly for advanced precancerous lesions (APL). Stool-derived host RNA biomarkers represent a promising approach to improve screening performance. However, numerous technical challenges are associated with extracting and quantifying this RNA, including an abundance of PCR inhibitors, a high microbial background, and varying levels of RNA degradation.

**METHODS:** We developed an extraction method for the isolation of host RNA from stool with improved sensitivity of detection and better diagnostic performance than the conventional phenol-chloroform-based extraction. ECB-Extract (<u>E</u>nrichment <u>C</u>apture <u>By</u> hybridization), uses hybridization capture of RNA under denaturing conditions by adding locked nucleic acid bases to the probes. The improved hybridization characteristics allow us to directly isolate host RNA from stool lysate.

**RESULTS:** ECB-Extract significantly improved PCR inhibitor removal, enabling scale-up of stool input and resulting in increased diagnostic sensitivity. We evaluated ECB-Extract against a conventional phenol-chloroform-based extraction method using the same panel of biomarkers in a pilot clinical cohort (N = 73). Using ECB-Extract, the sensitivity for detecting CRC and Advanced Precancerous Lesions (APL) was 96% and 50%, respectively, compared with 56% and 21% for the phenol-chloroform comparator at 96% specificity. In a larger cohort of 359 samples, ECB-Extract combined with a panel of the eight CRC-associated mRNA biomarkers achieved sensitivities of 95.7% for CRC and 51.2% for APL at a specificity greater than 90%.

**CONCLUSION:** These results demonstrate that ECB-Extract substantially improves stool host RNA extraction performance and, consequently, enhances diagnostic sensitivity for CRC and APL.

## Introduction

The gold standard method for the detection of Colorectal Cancer (CRC) and Advanced Precancerous Lesions (APL) is a colonoscopy. While reliable for detection of CRC and APL lesions, colonoscopy is an invasive and expensive procedure, requiring unpleasant preparation and missed days of work. Non-invasive stool-based tests are the most widely used alternatives to colonoscopy for colorectal cancer (CRC) screening. In the United States, FDA-approved stool-based screening options include fecal immunochemical tests (FIT)^1^, which detect occult blood in stool; Exact Sciences’s Cologuard, a multitarget stool DNA test combined with FIT^2^; and Geneoscopy’s ColoSense, which integrates multitarget stool mRNA detection with FIT^3^.

Host-derived RNA has emerged as a promising biomarker for colorectal cancer screening^4^. Because DNA copies per cell are limited, RNA targets can provide elevated signal through higher copy number and improve assay sensitivity, particularly important for detecting early-stage colorectal tumors and precancerous lesions.

Nucleic acid extraction is critical for molecular testing, as efficient removal of inhibitors directly determines assay sensitivity. However, methodologies for extracting host RNA from stool are far less developed than those for host stool DNA^5^. Most of the studies on stool host RNA use phenol–chloroform RNA extraction (Phenol Method)^6,7^ or magnetic silica bead based extraction (Silica Bead method)^8^. Some of these studies use commercial stool total RNA kits that were originally designed for the extraction of microbial RNA^9^. A key limitation of these commercial kits is that they do not enrich human RNA and have limited capacity for stool input. Overall, the major challenges for the isolation of human RNA from stool samples include the very low fraction of human RNA compared to gut microbial RNA. In addition, stool has high amounts of PCR inhibitors^10,11^ and contains abundant RNases, often resulting in stool-derived human RNA being partially degraded. The degradation levels vary among different samples.

Moreover, human cells can be shielded from lysis reagents by the ubiquitous mucus and cellular debris that are present in stool^12^. These challenges are further compounded by the extremely low copy number of RNA molecules derived from early-stage colorectal cancer and precancerous lesions.

Here, we describe ECB-Extract (Enrichment Capture By hybridization), a method designed to address these challenges. ECB-Extract can process arbitrarily large amounts of stool extract, concentrate host RNA, remove PCR inhibitors, and enrich both fragmented and intact RNA. The method incorporates several key innovations. First, locked nucleic acids (LNAs)^13^ are used in probe design to enhance binding affinity between probes and RNA; the increased stability of LNA base pairing enables efficient pulldown in the presence of high concentrations of chaotropic salts. Second, we developed a customized stool lysis and probe hybridization buffer with improved capacity to dissolve stool mucus and food debris while denaturing abundant RNases. Third, ECB-Extract employs both target-specific probes and poly(dT) oligos to capture RNA directly from crude lysates, enabling high capture efficiency for both fragmented and intact transcripts—an essential feature for efficient host RNA extraction from stool.

## Material and methods

### Stool RNA extraction

#### ECB-Extract Method

The ECB-Extract method employs proprietary technology developed by El Capitan Biosciences. Briefly, host RNA in stool is captured using a combination of poly(dT) and targeted single-stranded DNA probes immobilized on magnetic beads. Locked nucleic acids (LNAs) are incorporated into probe designs to enhance hybridization affinity and stability. A chaotropic agent is included in the lysis and binding buffer to disrupt stool mucus and food debris, facilitating efficient RNA release and capture.

#### Phenol Method

The E.Z.N.A. Stool RNA Kit (R6828, Omega Biotek) kit was used following the manufacturer’s protocol with minor modifications.

#### Silica Bead Method

The Dynabeads™ SILANE Viral NA Kit (37011D, Thermo Fisher Scientific) was used following the manufacturer’s protocol with minor modifications.

### Stool sample collection

Normal stool samples used for method development were purchased through a vendor in the US (Medix Biochemica).

Human clinical stool samples and associated clinical data were collected through a contract research organization (ZunTai medical technology, Guangzhou, China) from symptomatic and asymptomatic individuals scheduled to undergo colonoscopy. All participants were aged 43 years or older. Stool samples were collected at least 14 days prior to colonoscopy or at least 14 days following diagnostic colonoscopy and before any therapeutic or surgical intervention. The sample categorization and exclusion criteria is described in the Supplementary Information and Table S1. Participant demographic characteristics are summarized in Table S2. This study was approved by the ethics committees of all participating hospitals. Written informed consent was obtained from all participants prior to stool collection. The study was conducted in accordance with the Declaration of Helsinki and applicable local regulations.Participants self-collected fresh stool samples (15–40 g) into sealed fecal collection tubes. Samples were delivered to the hospital within 4 hours of collection and stored at −20 °C prior to processing. All stool samples collected in China were processed and analyzed locally at Guangdong Jiyin Biotech (Shenzhen, China).

### FIT testing

Clinical fecal immunochemical testing (FIT) was performed using Tellgen (Shanghai, China)

### RT-qPCR

RT–qPCR was performed to quantify transcript levels for each target gene using the OneStep PrimeScript™ III RT–qPCR 2× Master Mix (Takara Bio; part no. RR601), following the manufacturer’s instructions. Reactions were run on an ABI 7500 real-time PCR system (Applied Biosystems). Each 10 µL RT–qPCR reaction contained 4 µL of RNA eluate. Data acquisition and analysis were performed using Applied Biosystems software. Positive controls and no-template controls (NTCs) were included in all RT–qPCR runs.

### 5’:3’ Assay

RNA integrity was assessed using a modified 5′:3′ assay adapted from previously described two-step PCR methodologies.^14^ which are 2-step PCR assays. Briefly, extracted RNA was reverse-transcribed into cDNA using oligo(dT) primers and SuperScript IV Reverse Transcriptase (Invitrogen). Quantitative PCR (TaqMan™ Fast Advanced Master Mix, AppliedBiosystems) was then performed using one primer set targeting the 3′ end near the poly(A) tail and a second primer set targeting the 5′ region of the GAPDH transcript. RNA degradation was quantified by the ΔCq (5’Cq - 3’Cq), with ΔCq >1 indicating significant 5’ degradation.

### MS2 Assay

PCR inhibition in RNA eluates was assessed using MS2 bacteriophage as an external RNA spike-in control^15^. A fixed amount of MS2 bacteriophage RNA (Roche, part no. 10165948001) was added to an aliquot of each RNA eluate and, in parallel, to an equal volume of nuclease-free water. Reverse transcription quantitative PCR (RT-qPCR) targeting the MS2 RNA was performed on both reactions under identical conditions. PCR inhibition was quantified as the ΔCq value, calculated by subtracting the Cq value of the RNA eluate from that of the water control (Cq_{water} − Cq_{eluate}). Negative ΔCq values indicated the presence of PCR inhibitors in the RNA eluate.

### Data Analysis

Diagnostic performance (sensitivity and specificity) of the selected gene panel was evaluated using random forest models (sklearn module in python), with performance estimates obtained by averaging results from five-fold cross-validation repeated 10 times with different random data splits. Input features for the machine learning analyses were Cq values for each gene in the gene panel. For genes with undetectable expression in a given stool sample, the Cq value was set to 40.

## Results

### Comparison of PCR inhibitor levels in different stool host RNA extraction methods

High levels of PCR inhibitors pose a major challenge for stool extraction methods. Increasing sample input volume is one approach to improve the sensitivity of diagnostic assays for samples with low-copy-number of target RNA/DNA molecules.

However, with stool samples, increasing input often results in lower PCR signal (with higher PCR Ct values) due to the concomitant increase in PCR inhibitors. We evaluated whether ECB-Extract remains free of PCR inhibitors when scaling up stool input. We compared PCR Cq values across different RNA extraction methods using three genes measured from stool samples collected from three donors. One gene is a housekeeping gene (GAPDH), while the other two—TGFBI and HBA—are CRC–associated genes identified in our previous studies^16,17^. Compared with conventional stool host RNA extraction methods—the Phenol Method and Silica Bead method—ECB-Extract exhibited a linear decrease in PCR Cq values with increasing stool input (from 0.125g to 1g) in all the three stool samples (Figure 1a,b, c and Figure S1), whereas Cq values for the other two methods plateaued or increased in some of the stool samples, a characteristic signature of PCR inhibition. The effect of PCR inhibitors was further assessed by examining changes in Cq values (measured in GAPDH gene) as the volume of RNA eluate added to the PCR reaction was increased. For the ECB-Extract method, doubling the RNA template volume in PCR reactions gives the expected 1 Cq decrease in all the six samples tested (Figure 1d). For the Phenol Method and Silica Bead method, adding more volume of the RNA eluate negatively impacts the PCR signals (Cq increase) in most of the samples (Figure 1e,f). ECB-Extract consistently improved the Cqs with increased eluate input volume, likely because PCR inhibitors were being removed more completely.

**Figure 1:**
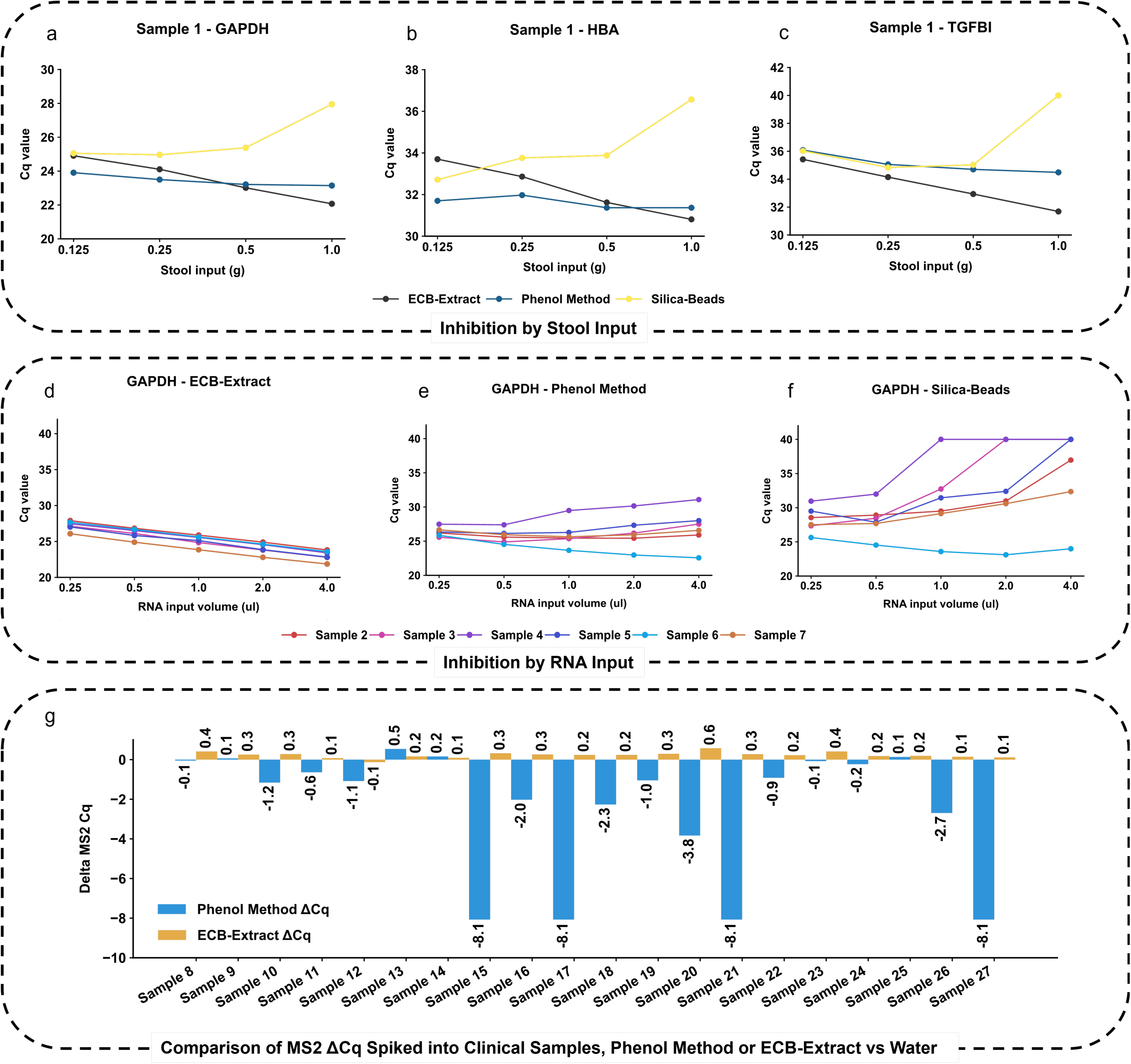
Comparison of residual PCR inhibitor levels across different stool host RNA extraction methods (ECB-extract, Phenol Method, Silica Bead method). a-c, The relationship between RT-PCR Cq value and the amount of stool input (0.125 g - 1g) measured in three host genes (GAPDH, HBA and TGFBI). A plateau or increase in Cq value with increasing stool input indicates the presence of PCR inhibitors in the extracted RNA. d-f, Relationship between the volume of host RNA template added to the RT–PCR reaction and the corresponding Cq values for the GAPDH gene across stool samples from six independent donors. A plateau or increase in Cq value with increasing RNA template volume is indicative of PCR inhibition. g,MS2 phage RNA was spiked in extracted RNA and water to measure the PCR inhibitor level. The difference (delta value) in MS2 Cq value measured by RT-PCR between the MS2 spiked into water controls and host RNA extracts is shown for 20 stool samples processed using ECB-extract and the Phenol Method.

One way to measure the PCR inhibitor level is to use MS2 bacteriophage as spiked-in RNA^15^. To further validate that the improvements in Cqs achieved by ECB-Extract are due to better inhibitor removal, we spiked MS2 phage into eluates from 20 clinical samples extracted by either the Phenol method or ECB-Extract. The MS2 ΔCqs (Cq of Water - Cq of Extract) show that many eluates from the Phenol Method still contain PCR inhibitors while eluates from ECB-Extract samples have no detectable PCR inhibitors (Figure 1g). In the ECB-Extract method, water controls exhibited higher Cq values than RNA eluates. Because MS2 RNA was present at very low concentration, the absence of background RNA in water controls likely promoted adsorption of MS2 molecules to PCR tube walls, resulting in lower effective template input and higher Cq values by RT-qPCR.

### Probe design strategy to capture stool host RNA with different levels of degradation

Host-derived RNA in stool comprises a mixture of fragmented and full-length transcripts. Fragmented RNA is presumed to arise from cellular debris and cell-free RNA, whereas full-length transcripts are more likely to originate from intact host cells. The degree of RNA fragmentation varies across clinical samples and is influenced by multiple factors, including sample collection and storage conditions, as well as inter-individual variability. To maximise recovery of host RNA from stool samples, magnetic beads were coupled with both poly(dT) oligonucleotides and target-specific capture probes. Poly(dT) oligonucleotides preferentially capture transcripts with intact 3′ poly(A) tails, whereas target-specific probes enable enrichment of fragmented transcripts that have lost their 3′ ends. Target-specific capture probes were designed to hybridize within or adjacent to the corresponding RT–qPCR assay regions for each gene of interest.

Where possible, probes were designed to span exon–exon junctions to minimize capture of genomic DNA from the same loci. Both the poly(dT) oligonucleotides and target-specific capture probes were modified with locked nucleic acids (LNAs) to enhance RNA–probe binding affinity in the presence of chaotropic agents. In a subsequent experiment, we evaluated five clinical stool samples and demonstrated that combining poly(dT) and target-specific capture probes enabled more efficient recovery of host RNA than either poly(dT)-only or target-specific capture approaches alone across samples with varying degrees of RNA fragmentation.

We first assessed RNA fragmentation in five clinical stool samples using a 5′:3′ assay (Materials and Methods). Two samples showed slight degradation (ΔCq < 1), whereas the remaining three samples exhibited severe degradation (ΔCq > 2) (Figure 2a). We next compared the performance of three bead-based capture strategies—poly(dT) beads, single-target capture beads, and combination beads incorporating both poly(dT) and target-specific capture probes on a single bead—for isolating GAPDH and TGFB1 transcripts. Poly(dT) beads and combination beads showed superior performance when mRNA degradation is minimal(Figure 2b,c). In contrast, samples with higher levels of RNA fragmentation were more efficiently captured using target-specific capture beads or the combination beads (Figure 2b,c). Overall, the combined poly(dT) and target-specific capture approach demonstrated the most consistent performance across stool samples containing a mixture of intact and fragmented RNA (Figure 2b,c).

**Figure 2:**
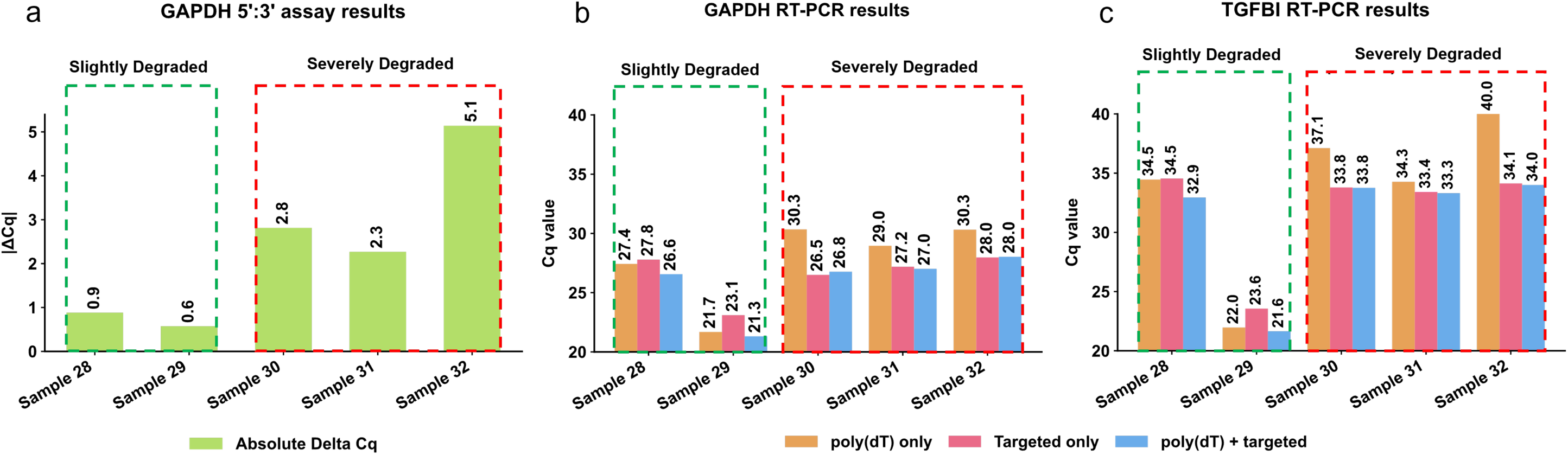
Combining poly(dT) and target capture probes improves host RNA extraction for samples with varying degrees of degradation. a, GAPDH 5’:3’ assay was used to access the degradation level for five stool samples. Samples with GAPDH 5’:3’ delta Cq < 1 were classified as ‘slightly degraded’, whereas the samples with GAPDH 5’:3’ delta Cq > 2 were classified as ‘severely degraded’. b, The RNA capture efficiency of three different hybridization probe designs (poly(dT), Target capture, and poly(dT) + target capture) was assessed by quantifying two host genes (GAPDH and TGFBI) using RT-PCR. The poly(dT) combined with targeted capture probes achieved consistent better results for both slighted and severely degraded samples.

### Comparison of Diagnostic Performance using Different Extraction Methods

To assess whether improved scalability with increased stool input translated into enhanced diagnostic performance, we compared the ECB-Extract method with a conventional Phenol Method in a cohort of 73 clinical stool samples (27 colorectal cancer (CRC), 21 advanced precancerous lesions (APL), and 25 controls comprising non-advanced precancerous lesions (NAPL) and colonoscopy-negative samples (NEG)). For the Phenol Method, 0.25 g of stool was used in accordance with the manufacturer’s recommendations. As shown in Figure 1a–c, increasing stool input beyond 0.25 g did not lead to further increases in RT–PCR signal using the Phenol Method. In contrast, the ECB-Extract method was performed using 1 g of stool input, as increased stool input resulted in a corresponding increase in RT–PCR signal.

We quantified the expression of 11 colorectal cancer (CRC)–associated genes (MMP7, TGFBI, HBA, MMP3, NKD2, IL11, PPBP, MYC, TCN1, CXCL8, and OLR1) that have previously been reported to be overexpressed in CRC stool samples^16^. For the ECB-Extract method, magnetic beads were functionalized with poly(dT) oligonucleotides in combination with target-specific capture probes corresponding to these 11 genes to enrich transcripts of interest. In contrast, the Phenol Method captures total RNA from stool without target-specific enrichment.

ECB-Extract consistently achieved higher sensitivity (at fixed specificity of 90%) than the conventional Phenol Method for each biomarker tested in both colorectal cancer (CRC) and advanced precancerous lesion (APL) detection (Figure 2a,d). When the 11 biomarkers were combined into a panel, the ECB-Extract method achieved a sensitivity of 96% for colorectal cancer (CRC) and 50% for advanced precancerous lesions (APL), whereas the Phenol method achieved a sensitivity of 56% for CRC and 21% for APL (Table 1, Figure 2 b,c,e,f).

**Table 1:** Diagnostic Performance Comparison; Phenol Method vs. ECB-Extract.

| Study | Test | CRC Sensitivity | Advanced Adenoma Sensitivity | CRC/Advanced Adenoma Specificity |
| --- | --- | --- | --- | --- |
| Sample Set (N = 73) | ECB-Extract | 96% | 50% | 96% |
|  | Phenol Method | 56% | 21% | 96% |

### Clinical Study for CRC and APL screening test

Given the improved sensitivity for colorectal cancer (CRC) and advanced precancerous lesion (APL) detection achieved with the newly developed stool host RNA extraction method, ECB-Extract, we proceeded to develop a CRC and APL screening assay and to evaluate its diagnostic performance in a larger cohort of 359 clinical samples (67 CRC, 58 APL, 77 non-advanced adenomas (NAPL), and 157 colonoscopy-negative controls (NEG)).

For the ECB-extract screening assay, we designed a panel comprising eight CRC-associated biomarkers (MMP7, TGFBI, NKD2, EPHX4, IL11, PPBP, IGF2, and HBA), together with a housekeeping gene (GAPDH) used for sample integrity and assay quality control. These eight CRC-associated genes were identified and validated in our previous study^16^. HBA represents the combined expression of the haemoglobin subunit genes HBA1 and HBA2. In prior work^17^, we demonstrated that haemoglobin mRNA can serve as an alternative to fecal immunochemical testing (FIT) for CRC and APL screening. To enable a head-to-head comparison with the conventional colorectal cancer (CRC) screening test, FIT was performed on a separate aliquot from the same stool samples.

The diagnostic performance of the ECB-extract screening assay was evaluated using a random forest machine learning model with five-fold cross-validation. The assay achieved sensitivities of 95.67% for colorectal cancer (CRC) and 51.21% for advanced precancerous lesions (APL). Specificity was 90.54% when assessed against samples with non-advanced adenomas (NAPL) or negative colonoscopy findings, and 93.83% when restricted to colonoscopy-negative samples alone (Table 2). In comparison, FIT achieved sensitivities of 80.15% for CRC and 17.41% for APL, with specificities of 95.48% for NAPL or colonoscopy-negative samples and 97.35% for colonoscopy-negative samples. Sensitivity across predefined clinical subgroups was evaluated and is summarized in Tables 3 and 4, including CRC stratified by stage, tumour size, and tumour location, as well as APL stratified by subtype, lesion size, and lesion location.

**Table 2:** Summary of the CRC Test Performance.

| **Colonoscopy/**<br>**Histopathology** | **N (359)** | **Clinical Performance**<br>**ECB-Extract** | **Clinical Performance**<br>**(FIT)** |
| --- | --- | --- | --- |
|  |  | **Sensitivity %** | **Sensitivity %** |
| **CRC** | **67**<br>**(18.66%)** | **95.67** | **80.15** |
| **APL** | **58**<br>**(16.16%)** | **51.21** | **17.41** |
| **CRC or APL** | **125**<br>**(34.82%)** | **75.04** | **51.04** |
|  |  | **Specificity %** | **Specificity %** |
| **NAPL or NEG** | **234**<br>**(65.18%)** | **90.54** | **95.48** |
| **NEG** | **157**<br>**(43.73%)** | **93.83** | **97.35** |

**Table 3:** Sensitivity of ECB-Extract screening assay and FIT for detecting different CRC and APL subgroups.

| Subgroup | N | ECB-Extract Sensitivity | FIT Sensitivity |
| --- | --- | --- | --- |
| **CRC Stage** |  |  |  |
| I | 7 | 84.30% | 37.10% |
| II | 14 | 100.00% | 85.70% |
| III | 22 | 95.50% | 79.10% |
| IV | 12 | 100.00% | 89.20% |
| Unknown | 12 | 93.30% | 91.70% |
| Stage I-III combined | 43 | 95.10% | 74.40% |
| **CRC tumor Size** |  |  |  |
| 10-19 mm | 3 | 73.30% | 43.30% |
| 20-29 mm | 6 | 81.70% | 21.70% |
| ≥ 30 mm | 28 | 100.00% | 87.90% |
| Unknown | 30 | 96.70% | 88.30% |
| **CRC tumor Location** |  |  |  |
| Proximal | 20 | 95.00% | 81.50% |
| Distal | 21 | 95.70% | 77.60% |
| Rectal | 26 | 96.20% | 81.20% |
| **APL Subtype** |  |  |  |
| Adenoma with carcinoma in situ/high grade dysplasia, any size | 12 | 63.30% | 25.80% |
| Tubulovillous adenoma, any size | 26 | 63.50% | 14.60% |
| Tubular adenoma ≥ 10 mm | 19 | 29.50% | 16.80% |
| Serrated lesion, ≥ 10 mm | 1 | 0.00% | 0.00% |
| **APL Lesion Location** |  |  |  |
| Proximal | 24 | 49.20% | 10.00% |
| Distal | 22 | 50.00% | 25.50% |
| Rectal | 12 | 57.50% | 17.50% |
| **APL Lesion size** |  |  |  |
| 5-9 mm | 1 | 100.00% | 20.00% |
| 10-19 mm | 32 | 41.90% | 11.60% |
| 20-29 mm | 12 | 73.30% | 25.80% |
| ≥ 30 mm | 10 | 35.00% | 9.00% |
| Unknown | 3 | 100.00% | 73.30% |

In the machine learning model, assay sensitivity and specificity can be adjusted by varying the classification threshold applied to the prediction probabilities. The specificity of the ECB-extract screening assay was initially set to match that of the FDA-approved multitarget stool DNA test^2^, whereas the specificity for FIT was set to reflect its reported specificity in the screening population^2^ (Table 2). For a more fair comparison with FIT, the threshold for the ECB-extract screening assay was further adjusted to achieve equivalent specificity to FIT. At matched specificity, the ECB-Extract based screening assay demonstrated higher sensitivity than FIT for both CRC detection (94.8% vs 80.5%) and APL detection (44.3% vs 17.4%) (Table S3). Consistent improvements in sensitivity with ECB-Extract based screening assay were observed across all evaluated CRC and APL subgroups at matched specificity (Table S4 and S5).

## Discussion

RNA-based assays have the potential to offer greater analytical sensitivity than DNA-based assays, as genomic DNA is present at two copies per cell, whereas transcript abundance can reach hundreds to thousands of RNA copies per cell, depending on gene expression levels. This inherent signal amplification at the RNA level may enhance detection sensitivity relative to DNA-based biomarkers, provided that RNA extraction methods consistently recover sufficient quantities of inhibitor-free host RNA. Improved RNA extraction efficiency therefore enables the effective use of lower-abundance RNA biomarkers while maintaining high diagnostic sensitivity and specificity. We developed ECB-Extract, an RNA isolation method that substantially improves extraction of human RNA from stool samples. Key improvements include enhanced removal of PCR inhibitors (Figure 1), scalability to larger stool input volumes (Figure1), enrichment of target mRNA from both fragmented and intact RNA species(Figure 2), and, ultimately, improved diagnostic performance compared to conventional stool RNA extraction method in clinical testing (Figure 3 and Table 1).

**Figure 3:**
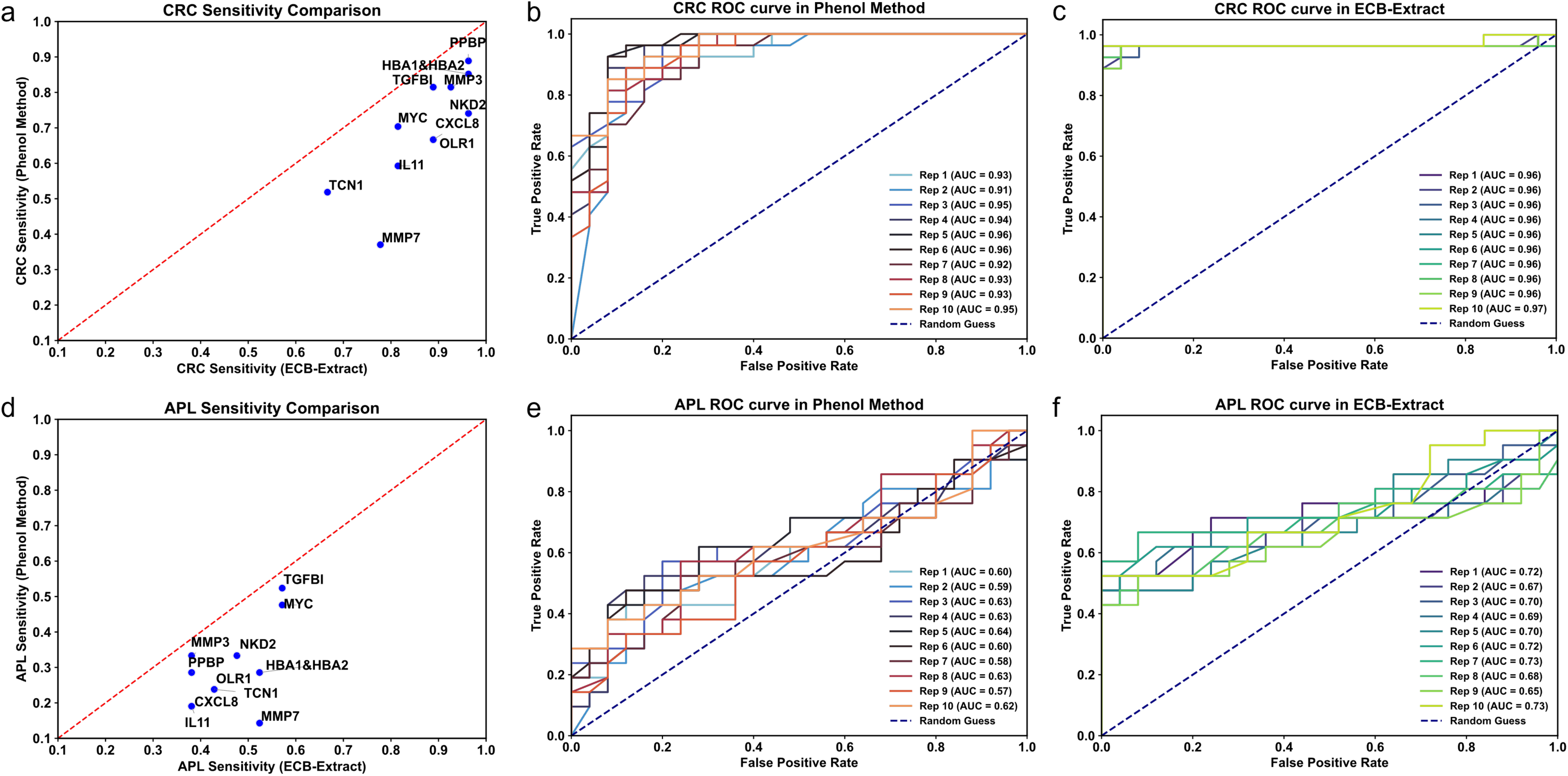
Comparison of the clinical performance between ECB-Extract and Phenol Method. a, CRC Sensitivity for each biomarker gene; Phenol Method (Y-axis) with ECB-Extract (X-axis). d, APL Sensitivity for Phenol Method (Y-axis) vs. ECB-Extract (X-axis). b,c,e,f, ROC curves for CRC or APL prediction from the machine learning model. Each line represents one random split of the training and test samples in the 5-fold cross validation; 10 repetitions were performed: (b) CRC prediction using the Phenol Method, (c) CRC prediction using the ECB-Extract method,(e) APL prediction using the Phenol Method, (f) APL prediction using the ECB-Extract method.

One innovation of ECB-Extract is the multiplexing of poly(A) selection and target-specific capture probes on a single magnetic bead. Poly(A) selection efficiently captures intact transcripts, whereas target-specific capture is more effective for fragmented RNA (Figure 2). For intact mRNA molecules, the poly(A) tail is often accessible, while internal target regions may be partially blocked by RNA secondary structure. In contrast, when transcripts are fragmented, target-specific capture outperforms poly(A) selection, resulting in improved recovery of degraded RNA species (Figure 2).

ECB-Extract allows capture of target RNA sequences in a concentrated chaotropic environment. Chaotropic salts, used in cell lysis, weaken hydrogen bonding. Hybridization-based capture of mRNA typically first requires bulk purification of nucleic acids. Using LNAs in our capture probes we can perform hybridization capture directly from the crude lysate, due to the stronger hydrogen bonding of LNA nucleotides.

Existing extraction techniques only isolate highly expressed stool mRNA biomarkers. This limits the pool of clinically useful RNA biomarkers. Many biomarkers, tested using ECB-Extract, showed substantially improved sensitivity with little loss of specificity. This boosted the number of clinically useful biomarkers.

Sensitivity for CRC detection was excellent using our 8-gene panel tested on 359 samples. CRC lesion detection was 95.67% (specificity of 90.54%, NAPL+NEG colonoscopy). The location and size of the lesion did not have a strong impact on detection rates, but detection of stage I lesions was lower than later stages (Table 3).

Consistent with previous report^2^, detection sensitivity for advanced precancerous lesions (APL) was higher for rectal lesions than for more proximal lesions, and lesions with high-grade dysplasia were detected with the highest sensitivity among APL subtypes (Table 4). The sensitivity of APL detection for all categories was 51.21% (specificity 90.54%). The sensitivity of detection for CRC/APL lesions compares favorably with previously published multitarget nucleic acid-based tests^2,3^. The sensitivity and specificity for FIT CRC/APL detection falls within the published performance ranges (Table 5)^3^. Compared with the performance of the mRNA panel, FIT was significantly worse overall, and across most clinical subgroups (Tables 3 and 4).

This study’s small sample size of clinical subcategories limits its impact. Some APL types had a limited number of samples, estimated diagnostic performance for these subtypes will be noisy. The CRC samples were enriched for rectal cancers relative to the expected incidence rates (Table 3). Rectal lesions may be easier to detect^2^, potentially inflating performance. However our detection rates for rectal CRC or distal lesions were similar (Table 3). Also later-stage cancers are comparatively over-represented vs. expected in an asymptomatic population (Table 3). Nevertheless, our detection sensitivity for stage I is comparable with the previously published multitarget nucleic acid-based test^2^ which has received FDA approval.

Our stool host RNA isolation method could improve non-invasive diagnostics/monitoring for general gastrointestinal (GI) disorders including; colorectal cancer screening, inflammatory bowel disease (IBD), GI infections and other GI-related diseases. Developing enhanced RNA-based stool tests for population screening is an important advancement for early detection of CRC and APL.

## Data Availability

All data produced in the present study are available upon reasonable request to the authors

## Acknowledgements

The authors would like to thank the following people for their contributions; Bowei Han, Youjia Ma and Yang Leng for data analysis; Ruikun Huang, Bingkun Huang, Wei Tang and Chaowei Fu for sample processing, Qing Zhou and Diana Cherbavaz for assay development.

## Author Contributions

Development of the original version of the ECB extract protocol: J.K., J.C., K.R, L.H., W.P. from El Capitan Biosciences; Development of the Python module for machine learning and data analysis pipeline: W.P. from El Capitan Biosciences; Optimization of the ECB extract protocol: H.L., D.C. and C.S. From Guandong Jiyin Biotech, Clinical sample validation; H.L., D.C and C.S. from Guandong Jiyin Biotech; Clinical data analysis and figure generation: Y.L. from Guangdong Jiyin Biotech. Drafting and reviewing of the manuscript: all authors.

## Funding

This research did not receive any specific grant from funding agencies in the public, commercial, or not-for-profit sectors.

## Notes

### Competing Interest Statement

The authors have declared no competing interest.

### Funding Statement

This study was funded by El Capitan Biosciences and Guangdong Jiyin Biotech.

### Author Declarations

Ethics committee/IRB of Foshan Nanhai District People's Hospital (Approval No. NYKY-2025-35-01), The Third Affiliated Hospital of Southern Medical University (Approval No.2025-伦审-001), Huizhou First hospital (KYLL-2025-044-01) and Jiangmen Central hospital (Approval No. 2024】228号A) gave ethical approval for this work.

