## Supplemental file for "A Novel Host Stool mRNA Extraction Method Improves Sensitivity of Detection for Colorectal Cancer and Precancerous Lesions"

**Study design of the clinical study:**

-	Clinical sample categorization:

Colonoscopy findings were classified into one of five categories: the Colorectal Cancer (CRC) group; individuals with colorectal cancer, stages I-IV. The Advanced Precancerous Lesions group: tubular or serrated adenomatous polyp ≥10 mm; ≥ 5 tubular or serrated adenomas; villous adenoma or tubulovillous adenoma with >25% villous component; or an adenoma with high-grade intraepithelial neoplasia (HGIN). Healthy Controls (HC) had normal colonoscopy findings. The Hyperplastic Polyp(s) group had <20 hyperplastic polyps. Finally, the Adenomatous Polyp(s) (NAPL) group: tubular adenoma(s) <10 mm in diameter, serrated adenomatous polyp(s) <10 mm, or ≥20 hyperplastic polyps.

-	Stool Sample exclusion criteria:

Exclusion criteria for the clinical sample collection; The following samples were excluded; specific diseases or conditions, such as inflammatory bowel disease (IBD), Celiac disease without dietary control, acute diarrhea, or gastrointestinal infections other than intestinal tuberculosis; Any other malignancy of the digestive system, including history of any malignancy diagnosed within the past 5 years, hereditary colorectal cancer syndromes (e.g., Hereditary Nonpolyposis Colorectal Cancer (HNPCC/Lynch Syndrome), Familial Adenomatous Polyposis (FAP); History of colorectal resection for any reason other than sigmoid diverticula or bowel resection >1 meter. Exclusion criteria for non-CRC control groups included individuals with lack of complete colonoscopy examination results and subjects who were pregnant or breastfeeding. This study was approved by the Ethics Committees of all hospitals at which samples were collected. Informed consent was given before stool was collected. The research was performed in accordance with the relevant guidelines and regulations, including the Declaration of Helsinki.

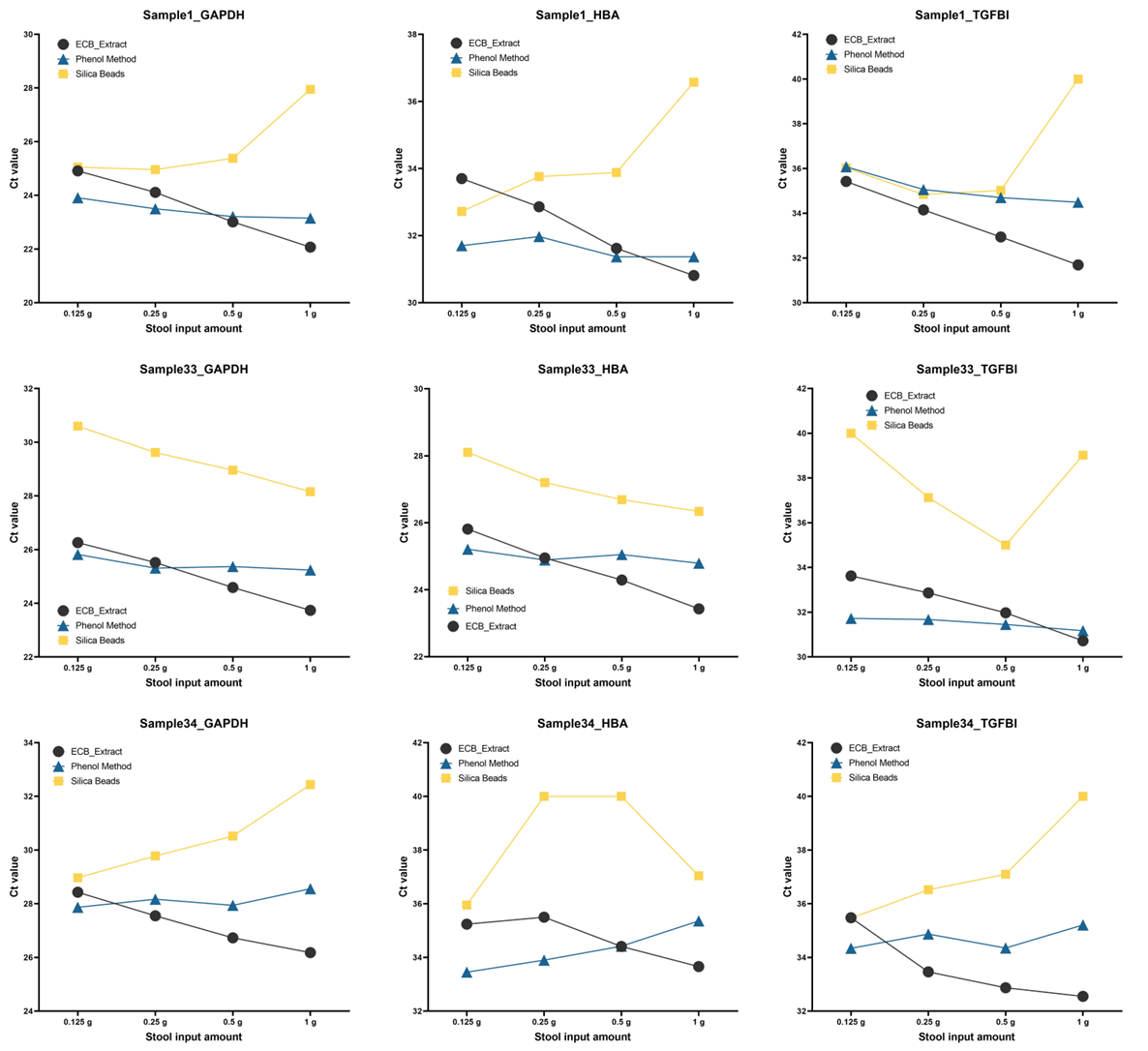

**Figure S1**.**RT-qPCR Cq values as a function of stool input across extraction methods.**

Cq values for GAPDH, HBA, and TGFBI were measured across increasing stool inputs (0.125–1.0 g) in three stool samples using ECB-Extract, phenol-chloroform, and silica bead extraction methods.

**Table S1: The definition of different sample labels in the clinical study.**

| **Group** | **Sub-group** |
| --- | --- |
| Colorectal cancer (CRC) | Stage I |
|  | Stage II |
|  | Stage III |
|  | Stage IV |
| Advanced precancerous lesions (APL) | Adenoma with carcinoma in situ/high grade dysplasia, any size |
|  | Serrated lesion, >= 10 mm |
|  | Tubular Adenoma, >= 10 mm |
|  | Tubulovillous adenoma, any size |
| Non-APL (NAPL) | 1-9 adenomas or sessile serrated lesions. <10 mm, non-advanced |
| NEG | Hyperplastic polyps or non-neoplastic lesions |
|  | No lesions on colonoscopy |

**Table S2: Baseline demographics for the ECB-extract CRC screening clinical study cohort**

| **Characteristic** | **Subgroup** | **Primary Effectiveness Population**  **(n = 359)** |
| --- | --- | --- |
| AGE | Mean (range) | 60 (45 years- 84 years) |
|  | 43-49 | 41 (11.42%) |
|  | 50-54 | 71 (19.78%) |
|  | 55-59 | 78 (21.73%) |
|  | 60-64 | 76 (21.17%) |
|  | >65 | 93 (25.91%) |
| SEX | Female | 204 (56.82%) |
|  | Male | 155 (43.18%) |

**Table S3: Overall Performance of ECB-Extract and FIT at matched specificity**

|  | **ECB Extract performance** | **FIT test performance** |
| --- | --- | --- |
| CRC sensitivity | 94.18% | 80.15% |
| APL sensitivity | 44.31% | 17.41% |
| Neg or NAPL specificity | 95.48% | 95.48% |
| Neg specificity | 97.65% | 97.35% |

**Table S4: CRC Sensitivity by Stage for ECB-Extract vs. FIT at matched Specificity**

| **Subgroup** | **ECB Extract Performance** | **FIT test Performance** |
| --- | --- | --- |
| **Stage I** | **71.40%** | **37.10%** |
| **Stage II** | **100%** | **85.70%** |
| **Stage III** | **95.50%** | **79.10%** |
| **Stage IV** | **100%** | **89.20%** |
| **Unknown stage** | **92.50%** | **91.70%** |
| **Stage I,II,III** | **92.50%** | **74.40%** |
| **Cancer Size** | | |
| **10-19 mm** | **70.00%** | **43.30%** |
| **20-29 mm** | **66.67%** | **21.70%** |
| **≥ 30** | **100%** | **87.90%** |
| **unknown size** | **96.67%** | **88.30%** |
| **Cancer Location** | | |
| **Distal** | **92.38%** | **81.50%** |
| **Proximal** | **95.00%** | **77.60%** |
| **Rectal** | **95.00%** | **81.20%** |

**Table S5: APL Sensitivity by Stage for ECB Extract vs. FIT at matched Specificity**

| **Subgroup** | **ECB Extract Performance** | **FIT test Performance** |
| --- | --- | --- |
| **Adenoma with carcinoma in situ/ high grade dysplasia, any size** | **57.50%** | **25.80%** |
| **Tubulovillous adenoma, any size** | **53.35%** | **14.60%** |
| **Tubular Adenoma, ≥ 10 mm** | **25.79%** | **16.80%** |
| **Serrated lesion, >= 10 mm** | **0.00%** | **0.00%** |
| **APL Location** |  |  |
| **Distal** | **42.73%** | **10.00%** |
| **Proximal** | **40.42%** | **25.50%** |
| **Rectal** | **55.00%** | **17.50%** |
| **Lesion size** |  |  |
| **5-9 mm** | **100%** | **20.00%** |
| **10-19 mm** | **34.38%** | **11.60%** |
| **20-29 mm** | **68.33%** | **25.80%** |
| **≥ 30 mm** | **25%** | **9.00%** |
| **unknown size** | **100%** | **73.30%** |
